# The effects of route previewing on adaptive gait and gaze behaviour under cognitive dual-task conditions in older adults

**DOI:** 10.64898/2026.10.02.26364602

**Authors:** Olivia McVeigh-Mellor, Adam J. Cocks, Liam O’Reilly, Toby Ellmers, Elmar Kal

## Abstract

**Background:** Safely walking through complex environments requires the use of visual information to plan and regulate steps, while managing competing cognitive demands. Age-related changes in visual control of gait and executive function may contribute to unsafe stepping behaviour, particularly under cognitive load. Route previewing may support visual-motor planning, but its effects under dual-task conditions in older adults are not well understood.

**Research Question:** Does cognitive dual-tasking influence gait and gaze behaviour during adaptive walking in older adults, and does previewing the walking route before gait initiation mitigate these effects?

**Methods:** Thirty-nine community-dwelling older adults (M_age_=73.4 years, SD=8.1; 14 male) completed four adaptive walking conditions, combining single- and dual-task walking with and without previewing. Per condition, participants completed six trials across different 8-m long coloured walking routes. Gait characteristics were assessed using an instrumented walkway and video-analysis, while gaze behaviour was recorded using mobile eye-tracking. Cognitive load was manipulated using a serial-subtraction task. Gait and gaze outcomes were evaluated using linear mixed-effects models.

**Results:** Dual-task performance significantly increased stepping errors, cross-steps, and double-limb support, while reducing stride length (*p* < .001). It also significantly increased the number and duration of task irrelevant fixations (*p* < .001). After previewing, participants exhibited more efficient gaze behaviour, as evidenced by looking further ahead (larger forward gaze transitions) and fewer irrelevant gaze fixations (*p* < .002). Significant Task-by-Preview interactions revealed that previewing mitigated the negative effects of dual-tasking on stepping errors and cross-steps (*p* < .001).

**Significance:** Route previewing prior to gait initiation attenuates negative effects of cognitive load on adaptive gait in older adults, likely by supporting visual-motor planning. These findings highlight the potential value of simple visual-planning interventions for improving adaptive gait in older adults.

## 1. Introduction

Falls in older adults are a significant cause of avoidable injury (Montero-Odasso et al., 2012) and often result in concerns about falling, activity avoidance and social isolation, which may ultimately contribute to further falls (Thomas et al., 2022; Ellmers et al., 2025). Falls are multifactorial and may occur due to intrinsic changes (including gait abnormalities, reduced strength, and comorbidities) as well as extrinsic environmental demands. Gait abnormalities are common in community-dwelling older adults (Verghese et al., 2005) and may impede the ability to adapt their gait to environmental constraints. For instance, older adults may be particularly challenged when safe movement requires rapid adaptations, such as avoiding a trip hazard or turning to avoid obstacles. Indeed, observational studies show that falls are often associated with an inability to safely shift bodyweight (Robinovitch et al., 2013).

Safely moving through complex environments requires the use of visual information to plan and regulate steps, while managing competing stability and attentional demands, especially in complex environments such as busy public spaces (Woollacott & Shumway-Cook, 2002; Al-Yahya et al., 2011). This can be particularly challenging for older adults, for whom balance becomes more reliant on attention-demanding conscious control, making their gait performance more vulnerable to disruption when attention is divided (e.g., cognitive dual-tasks [Boisgontier et al., 2013; Muhaidat et al., 2013; Muir et al., 2020; Li et al., 2018]). For instance, Cocks et al. (2025) showed that older adults make more “cross-steps” (defined as one foot crossing the midline of the body or across the lateral border of the other foot) while walking along a complex, winding route under cognitive dual-task conditions than under single task conditions. Such cross-steps narrow the base of support and may disrupt the safe controlled forward progression of the center of mass (Baker, 2018), making them a potentially unsafe stepping strategy (Cocks et al., 2025).

One particular way in which cognitive load may disrupt adaptive gait control is through changes in older adults’ gaze behaviour. During walking, we direct our gaze to simultaneously gather visual information about potential environmental hazards and plan upcoming steps accordingly, whilst also using vision to make online rapid adjustments to ongoing steps and ensure accurate foot placement (e.g., Reynolds & Day, 2005; Chapman & Hollands, 2006; Matthis et al., 2017). Indeed, it has been shown that high-risk older adults are more likely to shift their gaze away from a current stepping target before fully completing the step and to spend less time previewing more distant constraints or hazards on their route (Chapman & Hollands, 2006; Yamada et al., 2012). These gaze patterns are associated with slower gait and poorer stepping accuracy (Ellmers et al., 2016). Cognitive loading may compound these changes in gaze behaviour. For instance, cognitive dual-tasking has been shown to increase fixations on task-irrelevant areas and reduce time spent looking at stepping targets and other relevant route features in young adults (Russo et al., 2026). Additionally, Ellmers et al. (2016) reported that cognitive dual-tasking caused young adults to direct their gaze away from their walking path towards task-irrelevant areas, resulting in slower movement and reduced stepping accuracy. Similarly, Zukowski et al. (2020) found that older adults navigating distracting, real-world environments more often direct gaze toward distracting stimuli, accompanied by shorter fixation durations interpreted as reducing the acquisition of task-relevant visual information. They later found that cognitive dual-tasking shifted older adults gaze towards the near walking path and away from more distant environmental features (Zukowski et al., 2021). However, evidence directly examining the effects of cognitive dual-tasking on gaze behaviour and risky stepping during adaptive walking in older adults remains limited. Therefore, our first aim was to determine how cognitive dual-tasking affects gaze behaviour and the occurrence of risky stepping strategies during adaptive walking in older adults.

Cognitive loading may also reduce the visual information available for anticipatory motor planning (Zukowski et al. 2020; 2021; Ellmers et al., 2016). Ellmers et al. (2016) suggested that cognitive and visual demands compete for shared attentional and working memory resources and thereby reduce an individual’s ability to build and maintain a spatial representation of their route while walking. This may come at the expense of planning future stepping actions and increase reliance on immediate visual information to control/correct ongoing steps, something which has been associated with increased conscious control of gait; (Ellmers & Young, 2019). Therefore, especially whilst dual-tasking, the requirement to simultaneously allocate attention to cognitive, visual, and motor processes is challenged. Considering a limited-capacity account of attention, in which concurrent tasks share finite central processing resources (Tombu & Jolicœur, 2003; Repovš & Baddeley, 2006), this competition for resources may disrupt the acquisition and maintenance of visual information and adversely affect both cognitive-task performance and walking safety.

One potential strategy to overcome this limitation is route previewing. By allowing individuals to inspect upcoming stepping constraints before gait initiation, such previewing would thus support the formation of a spatial representation of the walking environment before attentional resources become divided. Curzon-Jones & Hollands (2018) demonstrated that visual previewing improves visuomotor control and stepping performance, suggesting that information acquired before movement can be used to support feedforward planning and spatial awareness reducing the need for continuous online visual search during walking. However, their study did not manipulate cognitive load. Therefore, the second aim of this study was to investigate if route previewing attenuated cognitive dual-task related changes in gaze and gait behaviour during adaptive walking in older adults.

Collectively, the literature suggests that successful adaptive walking depends on effective integration of visual sampling, attentional control, and motor planning. However, it remains unclear whether these gait changes can be mitigated by providing an opportunity to acquire information about the route before movement begins. Therefore, the present study aimed to examine changes in gait and gaze behaviour when attention is diverted from movement (single versus dual-task walking) and determine whether route previewing can reduce these negative effects by enhancing visual search prior to gait initiation.

### Hypotheses

H1 Cognitive dual-tasking will impair adaptive gait, resulting in increased stepping errors, cross-steps, and motor and cognitive dual-task costs.

H2 Cognitive dual-tasking will disrupt visual behaviour, resulting in more task-irrelevant and fewer task-relevant gaze fixations.

H3 Previewing the walking route before movement will attenuate these effects, resulting in improved gait performance, reduced motor and cognitive dual-task costs, and more task-relevant gaze behaviour.

## 2. Methods

### 2.1. Study design

This was a cross-sectional, within-subjects repeated-measures experimental study, whereby all participants completed four adaptive walking conditions comprising single-task and dual-task walking, with and without route previewing. The study protocol was approved by Brunel University of London’s College of Health, Medicine and Life Sciences’ Research Ethics Committee (reference: 45431-A-Sep/2024-52891-1).

### 2.2. Participants

An a-priori power analysis was conducted using G*Power version 3.1.9.7 (Faul et al., 2007). For a repeated-measures ANOVA with four measurements, assuming a medium effect (f = .25), α = .05, 95% power, the required sample size was 36 participants. Allowing approximately 10% for attrition or unusable data, a recruitment target of 40 participants was established. The anticipated medium to large effect of cognitive dual-tasking on cross-stepping behaviour was informed by preliminary findings from related work within the research group, subsequently published by Cocks et al. (2026).^1^

Participants were eligible if they were aged 60 years or older, had neither a musculoskeletal condition affecting balance nor progressive neurological diagnosis, were able to walk unaided for 10 meters, stand unsupported for at least 2 minutes, and had normal or corrected-to-normal vision. Participants were recruited through the Brunel Older People’s Reference Group – a group of community-dwelling older adults living across West London.

### 2.3. Procedures

All participants provided written informed consent prior to taking part in the study.

#### 2.3.1. Baseline assessments

Prior to undertaking the experimental protocol (see below), participants completed a battery of baseline measures related to their demographics, balance, and psychological and cognitive function. Concern (or “fears”) about falling was assessed using the Falls Efficacy Scale – International (FES-I; Yardley et al., 2005) and perceived control over falling was assessed using the Updated Perceived Control Over Falling Scale (UP-COF; Ellmers et al., 2023). General cognitive function was screened using the Montreal Cognitive Assessment (MoCA; Nasreddine et al., 2005). We also assessed habitual physical activity level (Physical Activity Scale for the Elderly; PASE; Washburn, 1993) and functional mobility (Timed-up-and-go; TUG; Podsiadlo & Richardson, 1991). Visual acuity was assessed using a Snellen chart. Level of education and self-reported number of falls in last 12 months were also recorded.

#### 2.3.2. Equipment

Figure 1 shows a visual summary of the experimental set-up, adapted from Yamada et al. (2011) and Cocks et al. (2025). Walking trials were conducted on an 8m instrumented walkway (Zeno Walkway system, ProtoKinetics, Havertown, PA, USA) with pre-defined routes displayed using coloured 10×10cm squares. The walkway allows automated assessment of gait parameters (see below for description of key parameters of interest). An integrated camera, positioned at the end of the walkway, recorded the walking trials to allow stepping errors and cross-steps to be assessed offline.

**Figure 1:**
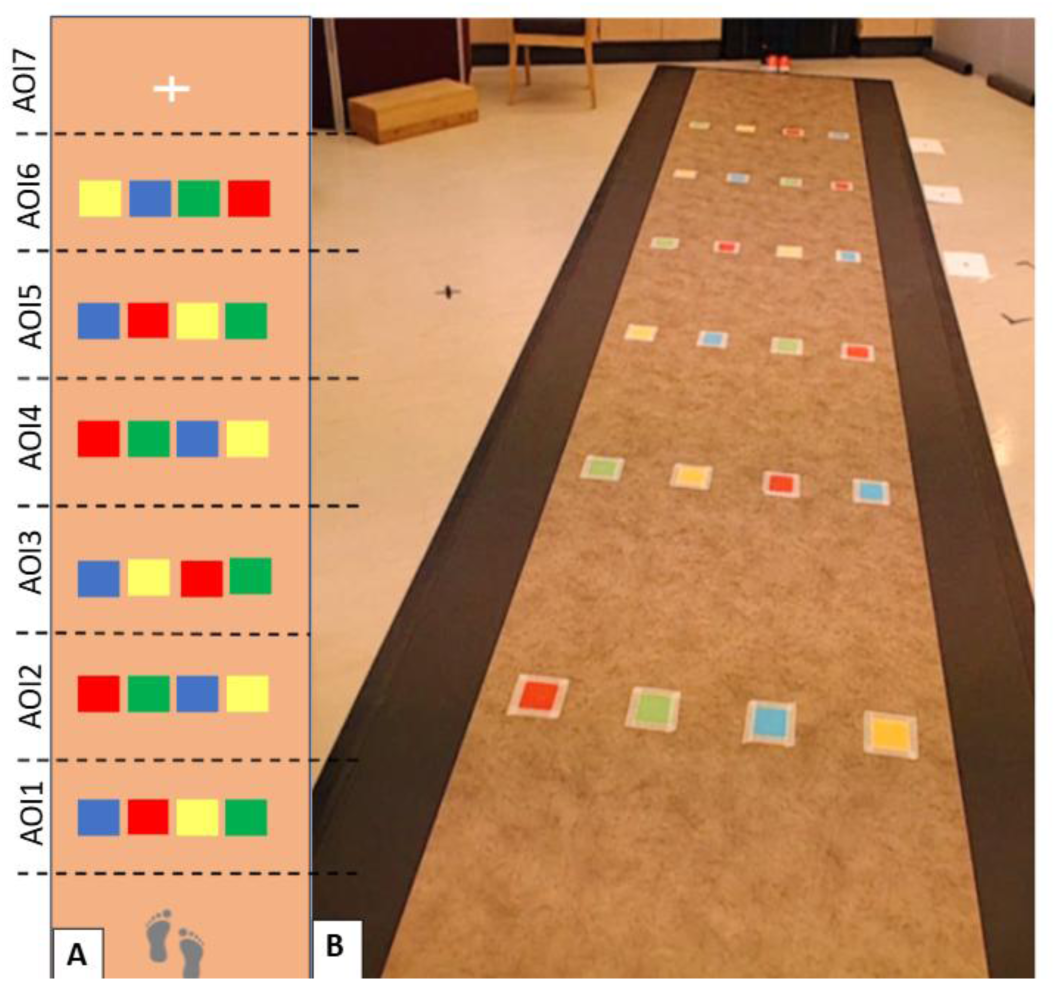
A) Visual schematic of the Zeno walkway with coloured routes and defined gaze areas of interest (AOI 1-7); B) reference image of the walkway from the video camera positioned at the end of the walkway used to assign cross-steps and stepping errors.

Participants wore SensoMotoric Instruments (SMI, Teltow, Germany) eye tracking glasses. Eye movements were sampled at 50 Hz, while a forward-facing scene camera on the nose bridge recorded the field of view at 25 frames per second (1920×1080 pixel resolution). The glasses were calibrated using 3 points prior to each walking condition and the calibration was then visually verified using a simple target check between walking trials and recalibrated if necessary.

#### 2.3.3. Experimental procedure

Participants completed the walking conditions in the following fixed order to ensure the previewing condition always came last to minimise carryover of route information acquired during deliberate previewing: (1) single-task – non-preview (STNP); (2) dual-task – non-preview (DTNP); (3) single-task – with preview (STP) and (4) dual-task – with preview (DTP). For each condition, all 4 coloured routes were used, with two routes used twice for a total of 6 trials per condition. All participants walked the same routes in a pre-set order but never walked the same-coloured route on consecutive trials.

The cognitive task required participants to verbally serially subtract in sevens, starting at a randomly determined three-digit number (Hausdorff et al., 2008). Following the final experimental condition, participants sat comfortably and carried out the serial subtracting task for 30 seconds (i.e., cognitive single-task condition). To allow for direct comparison of baseline cognitive task performance with cognitive performance during walking trials (which varied in duration), performance was expressed as the number of correct responses / second. This was obtained by dividing the correct number of responses in each trial by that trial’s duration (in seconds).

Prior to all trials, participants were positioned at the start of the walkway, facing away. For ST conditions, participants were given (i) the designated coloured route to follow, prior to (ii) a “Go” instruction after which they turned 180° and started walking along the designated coloured route. During the DT condition participants were instructed that trial’s designated colour route followed by the starting number for the serial subtracting task (participants would only start counting after receiving the “Go” signal). Finally, during the previewing conditions, once instructed which coloured route to follow, participants turned 180° to face the walkway and initiated a 10 second stationary previewing phase (timed using a stopwatch; based on Curzon-Jones and Hollands, 2018). Participants then turned away from the walkway again, waiting for the “Go” instruction to turn back and start walking in isolation (STP) or start walking whilst simultaneously serially subtracting (DTP only).

#### 2.3.4. Manipulation Checks

Between conditions participants had a short rest and completed a series of questionnaires. These were intended as manipulation checks, characterising participants’ self-reported levels of cognitive effort (The Rating Scale for Mental Effort, RSME; Zijlstra,1993), conscious movement processing (Movement-Specific Reinvestment Scale-state-version; Ellmers, & Young, 2018) and state anxiety (Mental Readiness Form, MRF; Krane, 1994). Together, these measures allowed us to verify whether the experimental manipulations influenced cognitive effort and attentional allocation as intended, while also allowing potential anxiety-related influences on gait performance to be identified and accounted for during interpretation of the results.

### 2.4. Outcome Variables

#### 2.4.1. Gait Outcomes

Mean gait speed (m/s), stride length (cm), and double limb support phase duration (% of the gait cycle) were calculated across all trials, using the Zeno Walkway Protokinetics software. For each trial, the final step was removed post-processing to exclude the participant’s transition off the walkway and ensure only the adaptive task was measured.

In addition, the following indicators of unsafe stepping strategies were obtained from analysis of video recordings (in line with Cocks et al., 2025): cross-steps (defined as one foot crossing the midline of the body or across the lateral border of the other foot) and stepping errors (stepping on an incorrect coloured route or missing a target completely). Identified cross-steps were cross-checked with the Zeno Walkway center of pressure output. Two researchers independently reviewed a selection of 16 participants videos for all four conditions, with perfect inter-rater reliability observed (100% agreement). We expressed cross-steps as a proportional measure (number of cross steps/total steps per trial) rather than a raw count to account for differences in number of steps between conditions and participants.

#### 2.4.2 Gaze Behaviour Outcomes

Gaze behaviour variables were assessed using the automatic event labelling capacity of the semantic gaze mapping mode of BeGaze software (V3.7 SensoMotoric Instruments, Boston, MA, 2018), which used a dispersion-based algorithm to identify fixations as groups of consecutive points within a maximum separation of 1 degree of the visual angle, and automated minimum fixation duration of 80ms (Salvucci & Goldberg, 2000).

Each gaze fixation was assigned to one of eight pre-defined AOIs in the participants’ field of view, and the duration of each fixation is measured in milliseconds. The first seven AOIs (Figure 1.A) were task-relevant areas of the walkway. The eighth AOI was assigned to any fixations occurring outside the walkway, all of which were considered irrelevant areas of the scene (e.g., windows and ceiling). The seven task-relevant AOIs were chosen to divide the walkway by the pathway targets, allowing assessment of gaze transitions defined as a fixation change between AOIs during walking. Unassigned fixations (where the crosshair was not visible) were counted for each trial. Two raters categorized 3 walking trials from two participants and compared results as a calibration procedure to maximize the consistency of fixation categorization across participants and trials, each rater then processed 50% of the data.

For each condition we assessed mean area and duration of first fixation (Ellmers & Young, 2019), mean number of task-relevant transitions and fixations (Ellmers et al, 2016), number of irrelevant transitions (whereby the transition was away from the walkway towards task irrelevant areas e.g. window) and fixations, and average task-relevant and irrelevant fixation duration during walking (Elmers et al, 2016; Walsh & Snowball, 2023). The first fixation (at start of each trial) was defined as the first fixation where participants could see the whole length of the walkway after turning. The number of gaze fixations per trial was defined as the number of fixations from the first step towards the walking targets to when the final target went out of view.

Maximum forward transition size was used to quantify the largest single forward shift in gaze along the walkway during walking. For this outcome, the walkway was divided into six sequential areas of interest (AOIs) (adapted from Zukowski et al., 2021), ordered according to their position along the route (AOI 1 = nearest and AOI 6 = furthest). For each pair of consecutive fixations, the forward transition was calculated as the difference between the AOI numbers. For example, a shift from AOI 1 to AOI 4 had a transition magnitude of three AOIs. The largest forward transition observed during each trial was retained, with larger values indicating a greater forward shift of gaze towards more distant areas of the upcoming walking route. This measure was included to characterise the extent of forward visual exploration during walking, reflecting how far gaze was shifted towards upcoming areas of the route in a single transition.

#### 2.4.3. Overall Motor and Cognitive dual-task performance (DTC)

We quantified dual-task costs as the percentage change in performance from single- to dual-task walking, using the following equation (Montero-Odasso et al., 2012):

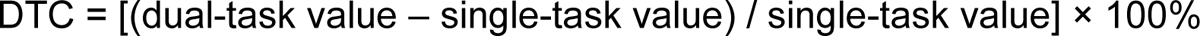

DTCs were calculated both for walking speed (DTC_sp) and stride length (DTC_sl) and the cognitive task (DTC_cog). Please note that we adjusted the general equation above, such that for all measures a positive DTC reflects a *reduction* in performance for dual-versus single-task performance.

#### 2.4.4. Data exclusion

Walkway data was removed at trial level if issues were noted during data collection (e.g., recording start failed or participant stopped walking). For both the STNP and DTNP conditions, 1/6 trials were excluded for three participants, and 2/6 trials for one participant. For the STP condition, 1/6 trials from two participants were excluded, while for the DTP condition, 1/6 from one participant was excluded.

Gaze behaviour data was excluded if extreme or unassigned values were noted that were clearly inaccurate/due to measurement error (e.g., poor eye tracker calibration resulting in infeasibly low numbers of fixations). The percentage of frames without position data per trial was calculated, and trials were excluded if 50% or more were unassigned (similar to Ellmers et al., 2016). If ≥3/6 trials for a single condition were excluded due to poor quality, the whole condition was excluded. Three participants were entirely excluded from the eye tracking analysis due to poor data quality. In total, 4 participants were removed from the STNP, DTNP and STP conditions, and 6 were excluded from the STP condition of the 936 trials collected, 48 (5.1%) were excluded due to missing or unusable data, leaving 888 trials (94.9%) for analysis.

### 2.5. Statistical Analysis

All statistical analyses were conducted using jamovi (Version 2.6) with the GAMLj mixed modelling module. Statistical significance was set at p < .05.

Descriptive statistics were calculated for all gait variables, gaze behaviour measures, and cognitive dual-task cost variables. Linear mixed-effects models were fitted using restricted maximum likelihood estimation with participant included as a random intercept to account for within-subject repeated measures. This also ensured all available data for each participant was used regardless of whether a participant had missing data for one or more conditions. Experimental condition was modelled using two fixed factors: Task (single-task, dual-task) and Preview (no-preview, preview), together with the Task × Preview interaction. This allowed us to examine the independent effects of cognitive loading and route previewing, as well as their interaction.

Walking speed was included as a covariate in the analyses of stride length, cross-step proportion, stepping errors, total double support, number of relevant transitions, number of irrelevant fixations, and maximum forward transition size (to control for the potential confounding influence of walking speed on gait and visual-search measures expected between conditions). Significant interaction effects were followed up by pairwise comparisons of estimated marginal means (EMM) and associated 95% confidence intervals were calculated for all conditions.

Cognitive dual-task cost was analysed separately from the factorial models because it represents the relative change between baseline and dual-task cognitive performance and therefore could not be modelled within the Task × Preview factorial design. A linear mixed-effects model was fitted with Preview (no-preview, preview) entered as a fixed effect and participant included as a random intercept.

The self-report manipulation-check measures (mental effort, state anxiety, and conscious movement processing) were analysed separately using 2 × 2 repeated-measures ANOVAs with Task (single-task, dual-task) and Preview (no-preview, preview) as within-subject factors.

## 3. Results

### 3.1 Participants

Forty-one healthy older adults consented to participate in the study. However, two participants were excluded due to low MoCA scores (<22) (Carson et al., 2018). Characteristics of the final sample of 39 participants are summarised in Table 1.

**Table 1:** Participant characteristics (N = 39)

| Demographics | Mean (SD) | Range |
| --- | --- | --- |
| Age | 73.4 (8.1) | 60-87 |
| Sex (Male/Female) | 14/25 | n/a |
| Height (cm) | 165.0 (12.3) | 125-185 |
| Weight (kg) | 76.5 (15.5) | 43-110 |
| Visual Acuity* | 20/25 (20/20–20/25) | 20/20-20/40 |
| Timed-up-and-Go (seconds) | 7.8 (1.6) | 6-11 |
| Montreal Cognitive Assessment (0-30) | 26.7 (2.3) | 23-30 |
| Falls Efficacy Scale - International (16-64) | 23 (6.1) | 16-42 |
| Number of participants with fall in previous year (N; %) | 9 (23%) | n/a |
| Updated Perceived Control over Falling Scale (0-20) | 17.3 (2.63) | 12-20 |
| Education (High school/University/post-grad/other) (N) | 13/16/4/4 | n/a |
| Physical activity (high/medium/low) (N) | 10/22/7 | n/a |

Note \* Median and IQR values presented

### 3.2 Gait outcomes

Gait outcomes are summarised in Figure 2. Participants walked significantly slower during dual-task when compared to single-task conditions, F_Task_ (1,114) = 112.73, p < .001, and walked significantly faster following previewing when compared to no-preview conditions, F_Preview_ (1,114) = 24.91, p < .001. The Task x Preview interaction was found to be non-significant, F_TaskxPreview_ (1,114) = 1.51, p = .222.

**Figure 2.**
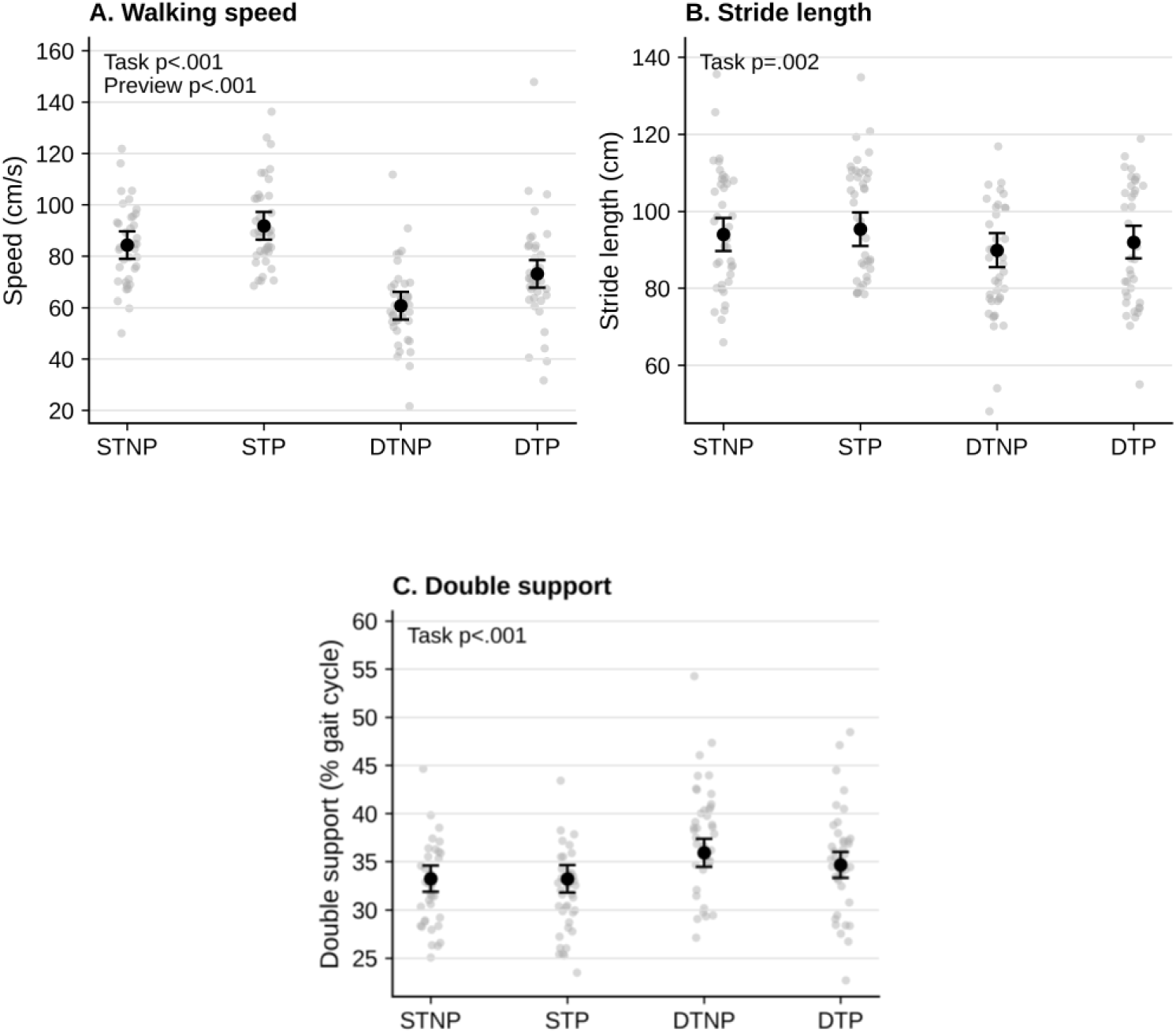
Effect of previewing and dual-tasking on gait outcomes during adaptive walking. Outcomes include: (A) Speed; (B) Stride length; (C) Double Support; These are presented across the following conditions: single-task no-preview (STNP), single-task preview (STP), dual-task no-preview (DTNP), and dual-task preview (DTP). Grey points represent individual participant values. Black points represent group means with 95% confidence intervals and significant post-hoc comparisons are highlighted based on the LMM post-hoc pair-wise tests. Unless otherwise stated, values reported for pairwise comparisons represent estimated marginal means (EMM) derived from the fitted mixed-effects models.

Stride length was significantly shorter during dual-task walking, F_Task_ (1,118) = 10.28, p = .002. There was no significant main effect of previewing, F_Preview_ (1,114) = 3.60, p = .060, nor a significant interaction, F_TaskxPreview_ (1,112) = 0.19, p = .668.

Finally, total double limb support percentage was significantly greater during dual-task walking, F_Task_ (1,129) = 13.68, p < .001; however, no significant Preview or interaction effects were observed, F_Preview_ (1,118) = 2.06, p = .154, F_TaskxPreview_ (1,112) = 2.21, p = .140.

### 3.3. Stepping outcomes

Cross-step proportion was significantly higher during dual-task walking in comparison to single-task walking, F_Task_ (1,134) = 6.00, p = .016, but significantly lower following previewing when compared to no previewing, F_Preview_ (1,120) = 5.15, p = .025. The significant interaction effect, F_TaskxPreview_ (1,113) = 17.30, p < .001, indicated that previewing significantly reduced cross-step proportion during dual-task walking (EMMs: DTNP= 0.099, DTP = 0.041, p < .001), but not during single-task walking (EMMs: STNP = 0.035, STP = 0.0518, p = 0.628.

Participants made significantly more stepping errors during dual-task in comparison to single-task walking, F_Task_ (1,146) = 23.60, p < .001, and significantly fewer errors following previewing when compared to no previewing, F_Preview_ (1,125) = 17.83, p < .001. The significant interaction, F_TaskxPreview_ (1,114) = 28.91, p < .001, showed that previewing significantly reduced stepping errors during dual-task walking, (EMMs: DTNP = 0.84, DTP = 0.11, p < .001), but not during single-task walking (EMMs: STNP= 0.0, STP = 0.06, p = 0.909).

### 3.4. Gaze behaviour

Participants made significantly more task-irrelevant fixations during dual-task than single-task walking, F_Task_(1,115.8) = 10.40, p = .002, and significantly fewer irrelevant fixations following previewing, F_Preview_(1,102.8) = 5.83, p = .018. The Task × Preview interaction was not significant, F_Task×Preview_(1,113.4) = 2.53, p = .116. Similarly, irrelevant transitions away from the walkway were significantly greater during dual-task walking compared to single-task, F_Task_(1,116.5) = 10.99, p = .001. However, although previewing reduced transitions to task irrelevant areas the effect of previewing, F_Preview_(1,104.2) = 2.13, p = .148, and the interaction, F_Task×Preview_(1,113.2) = 1.24, p = .268, was not significant. Average irrelevant fixation duration was also significantly greater during dual-task walking compared to single-task, F_Task_(1,96.5) = 21.44, p < .001, and significantly lower following previewing (Figure 3A), F_Preview_(1,97.6) = 4.59, p = .035, with no significant interaction, F_Task×Preview_(1,94.4) = 2.40, p = .123.

**Figure 3.**
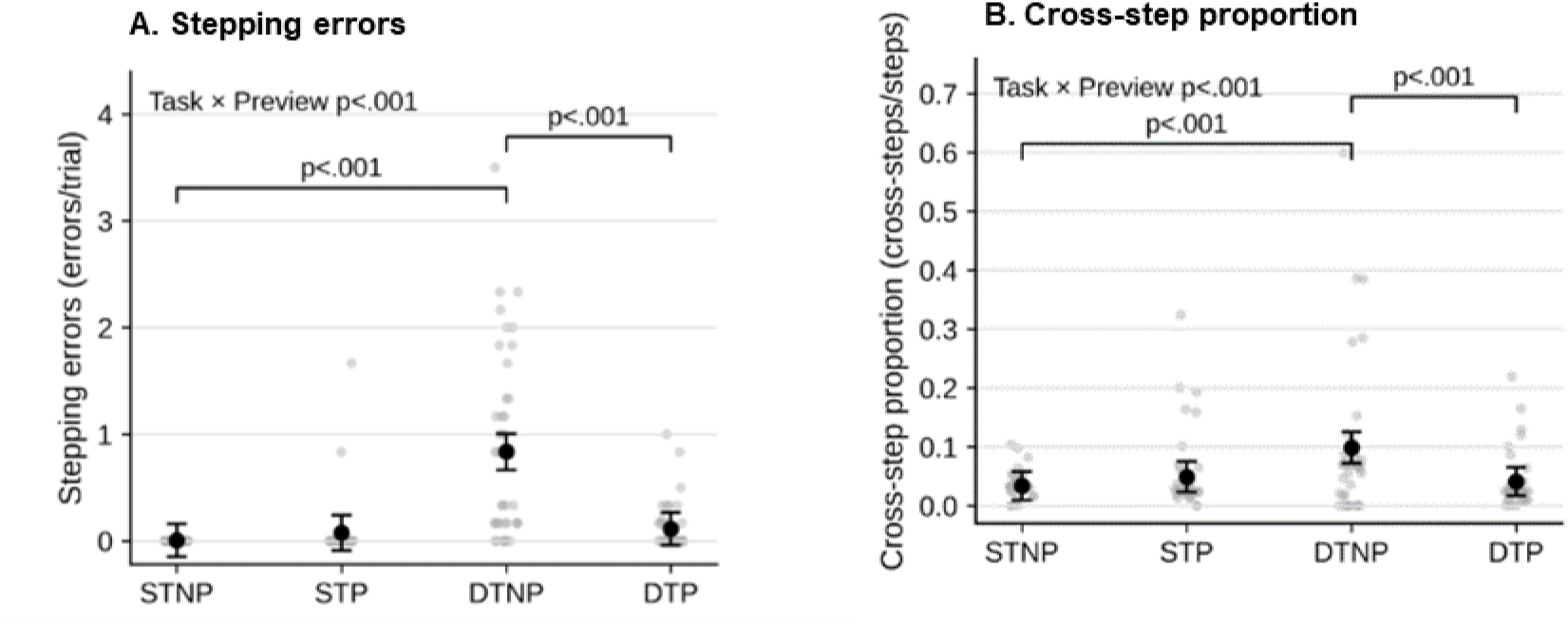
Effect of previewing and dual-tasking on stepping outcomes during adaptive walking. (A) Stepping errors and (B) Cross-step proportion; These are presented across the following conditions: single-task no-preview (STNP), single-task preview (STP), dual-task no-preview (DTNP), and dual-task preview (DTP). Grey points represent individual participant values. Black points represent group means with 95% confidence intervals and significant post-hoc comparisons are highlighted based on the LMM post-hoc pair-wise tests. Unless otherwise stated, values reported for pairwise comparisons represent estimated marginal means derived from the fitted mixed-effects models.

**Figure 3.**
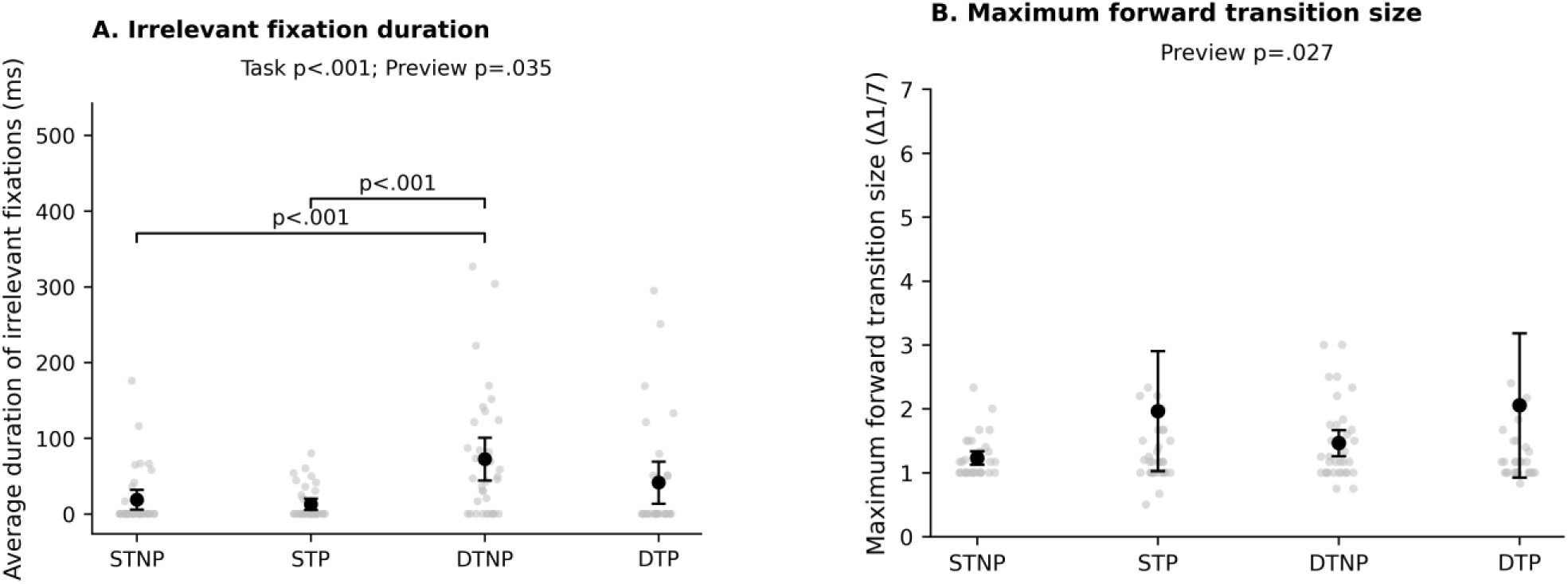
Gaze behaviour results. Panel A shows the average duration of irrelevant fixations across task and preview conditions. There were significant main effects of task (p < .001) and previewing (p = .035), with longer irrelevant fixations during dual-task compared with single-task walking and shorter irrelevant fixations following previewing. No significant Task × Preview interaction was observed. Significant Tukey-adjusted pairwise comparisons are indicated on the figure. Panel B shows the maximum forward transition size across task and preview conditions. Maximum forward transition size represents the largest forward transition between areas of interest (Δ1/7). There was a significant main effect of previewing (p = .027), with larger forward transitions in preview compared with no-preview conditions. No significant main effect of task or Task × Preview interaction was observed. Grey points represent individual participant values. Black points represent group means with 95% confidence intervals.

**Figure 4.**
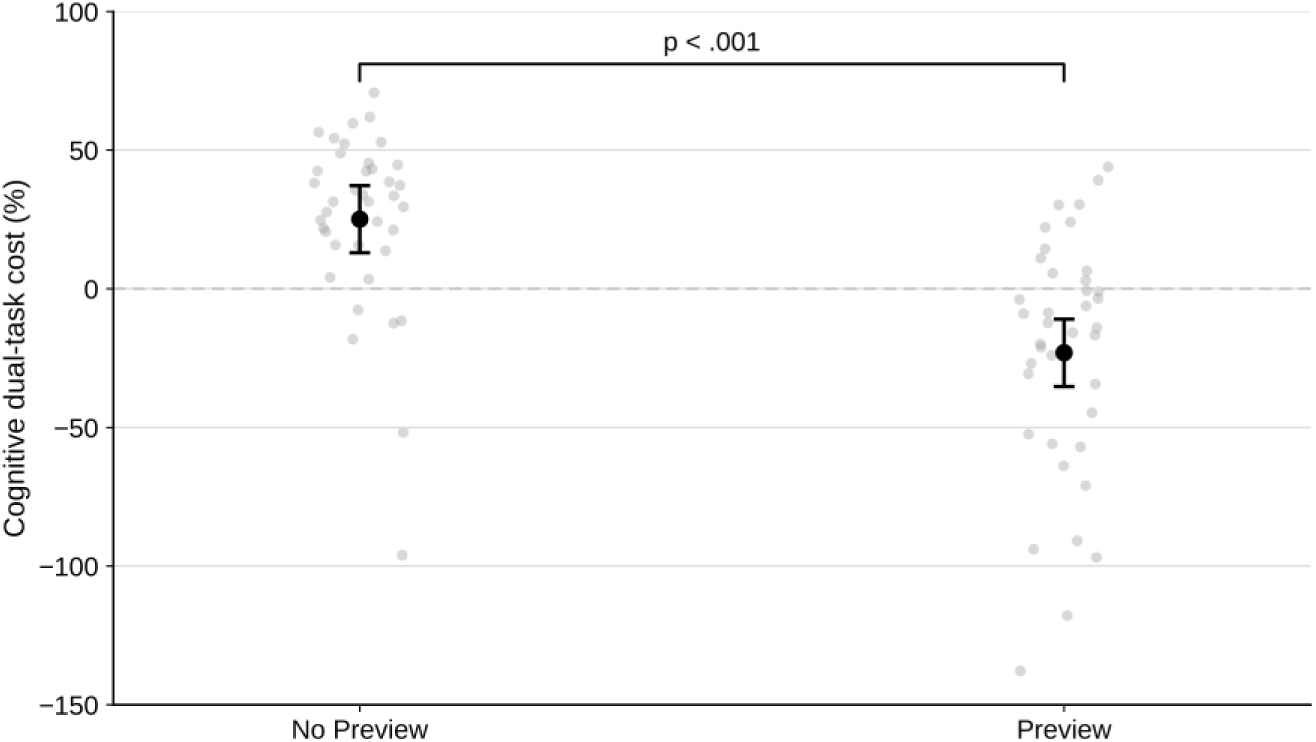
Cognitive DTC in no-preview and preview conditions. Grey dots represent individual participant values. Black dots represent group means with 95% confidence intervals. Negative DTC values indicate improved performance in dual-task compared to single-task conditions.

Measures relating to task-relevant visual planning showed limited effects of dual-tasking. First fixation location significantly differed by Task, F_Task_(1,96.5) = 7.71, p = .007, dual-tasking was associated with more proximal first fixation locations, with lower fixation-area scores observed in both dual-task conditions (DTNP = 1.10; DTP = 1.22) compared with the corresponding single-task conditions (STNP = 1.30; STP = 1.34); while neither Preview, F_Preview_(1,94.4) = 1.78, p = .185, nor the Task × Preview interaction, F_Task×Preview_(1,94.5) = 0.52, p = .472, were significant. Maximum forward transition size was significantly greater following previewing (Figure 3B), F_Preview_(1,105.9) = 5.00, p = .027, but was not affected by Task, F_Task_(1,117.1) = 0.03, p = .868, nor was the interaction effect significant F_Task×Preview_(1,112.5) = 0.04, p = .845. No significant effects of Task, Preview, or their interaction were observed for average task-relevant fixation duration or the number of task-relevant transitions (all p > .12; Table 2).

**Table 2:** Descriptive gaze outcomes across conditions, presented as Mean (SD), with significance (*p*) values reported (\**p* < .05, \*\**p* < .01, \*\*\**p* < .001)

| Outcome | STNP | STP | DTNP | DTP | Task p | Preview p | Task ×<br>Preview p |
| --- | --- | --- | --- | --- | --- | --- | --- |
| Area of First Fixation (1-7) | 1.30 (0.44) | 1.34 (0.46) | 1.10 (0.27) | 1.22 (0.39) | 0.007** | 0.185 | 0.472 |
| Number of task-relevant Fixations (n) | 24.1 (4.56) | 22.4 (5.34) | 31.2 (10.6) | 25.2 (6.64) | 0.002** | 0.018* | 0.116 |
| Number of Irrelevant Fixations (n) | 0.281 (0.747) | 0.112 (0.262) | 1.11 (1.60) | 0.486 (1.12) | 0.002** | 0.018* | 0.116 |
| Number of task-relevant Transitions (n) | 8.55 (2.42) | 8.75 (2.42) | 9.32 (4.32) | 8.08 (2.55) | 0.172 | 0.709 | 0.121 |
| Number of Irrelevant Transitions (n) | 0.157 (0.365) | 0.113 (0.224) | 0.577 (0.751) | 0.360 (0.713) | <.001*** | 0.148 | 0.268 |
| Average Duration of task-relevant Fixations (ms) | 266 (59.0) | 266 (58.3) | 255 (44.7) | 268 (58.4) | 0.511 | 0.483 | 0.393 |
| Average Duration of Irrelevant Fixations (ms) | 18.8 (39.1) | 13.0 (22.4) | 72.6 (84.6) | 41.5 (77.7) | <.001*** | 0.035* | 0.123 |
| Maximum Forward Transition Size ($\Delta 1/7$) | 1.23 (0.315) | 1.96 (2.79) | 1.47 (0.609) | 2.05 (3.21) | 0.868 | 0.027* | 0.845 |

Together, these findings suggest that dual-tasking disrupted visual attention primarily by increasing task-irrelevant gaze behaviour. Previewing partially reversed these effects by reducing task-irrelevant gaze fixations and promoting forward visual exploration. The consistent absence of Task × Preview interactions across gaze measures suggests that these benefits occurred similarly under both single and dual-task conditions, suggesting that previewing was advantageous irrespective of concurrent cognitive load.

### 3.4 The effect of previewing on cognitive and motor dual-task costs

Cognitive DTC was significantly lower in preview compared with non-preview conditions, F_Preview_ (1, 39.2) = 71.2, p < .001. Estimated marginal means indicated that participants demonstrated a cognitive DTC of +25.1% in no-preview condition (DTNP), compared with a cognitive dual-task of −23.1% (DTP) after previewing, representing a mean reduction of 48.2%. Motor (speed) dual-task cost was also significantly reduced following previewing, F(1,76) = 71.50, p < .001, with costs decreasing from M = 25.1% in the no-preview condition to M = −23.1% in the preview condition.

### 3.5 Manipulation Checks

Manipulation checks (Table 3) confirmed that the cognitive loading manipulation increased perceived mental effort. Mental effort was significantly greater during dual-compared with single-task walking, F_Task_ (1,38) = 91.53, *p* < .001, and significantly lower following previewing, F_Preview_ (1,38) = 12.24, *p* = .001. A significant Task × Preview interaction was also observed, F_TaskXPreview_ (1,38) = 4.58, *p* = .039, reflecting a smaller increase in mental effort from single- to dual-task walking following previewing. State anxiety scores were significantly greater during dual-task walking, F_Task_ (1,38) = 22.42, *p* < .001, and lower following previewing, F_Preview_ (1,38) = 21.52, *p* < .001. The Task × Preview interaction was also significant, F_TaskXPreview_ (1,38) = 6.50, *p* = .015, similarly reflecting a smaller increase in state anxiety scores from single- to dual-task walking following previewing. Conscious movement processing was significantly lower following previewing, F_Preview_ (1,38) = 21.73, *p* < .001, but was not affected by Task, F_Task_ (1,38) = 0.00, *p* = .950. However, there was a significant Task × Preview interaction, F_TaskXPreview_ (1,38) = 5.01, *p* = .031, reflecting a decrease in conscious movement processing from single- to dual-task walking without previewing, compared with an increase following previewing.

**Table 3:** Descriptive statistics and repeated-measures ANOVA results for manipulation-check measures across Task and Preview conditions.

| Condition / Effect | State Anxiety Mean (SD) | Conscious Movement Processing Mean (SD) | Mental Effort (RSME) Mean (SD) |
| --- | --- | --- | --- |
| STNP | 19.49 (26.30) | 2.26 (1.80) | 6.28 (9.01) |
| DTNP | 55.33 (58.40) | 1.87 (1.98) | 51.38 (30.39) |
| STP | 10.38 (18.97) | 1.15 (1.46) | 5.00 (6.69) |
| DTP | 31.18 (39.36) | 1.51 (1.70) | 40.23 (27.45) |
| **ANOVA** |  |  |  |
| Task | 22.42*** (.371) | 0.00 (.000) | 91.53*** (.707) |
| Preview | 21.52*** (.362) | 21.73*** (.364) | 12.24** (.244) |
| Task × Preview | 6.50* (.146) | 5.01* (.117) | 4.58* (.108) |

Note; State Anxiety is reported as a percentage, Conscious movement processing is scored from 0-16, and RSME 0-150. Values are mean (SD). ANOVA cells report F statistics with partial $\eta^2$ in brackets. \* $p < .05$ , \*\* $p < .01$ , \*\*\* $p < .001$ . Task = single- vs dual-task; Preview = no preview vs preview.

## 4. Discussion

Consistent with the study hypotheses, cognitive dual-tasking negatively affected adaptive walking performance, resulting in increased cross-steps and stepping errors in the absence of previewing (H1). Cognitive loading increased irrelevant fixations (H2) but otherwise did not significantly impact gaze behaviour during gait. Finally, previewing influenced gait stepping outcomes during both single-task and dual-tasking; however, H3 was not explicitly supported across all gait outcomes. Similarly, there was a significant overall effect of previewing on some gaze behaviour measures (number and duration of irrelevant fixations) though there was no evidence that previewing particularly altered gaze behaviour during dual-task conditions.

### 4.1 Dual-tasking and adaptive walking performance

The present findings support the hypothesis that cognitive dual-tasking alters gaze behaviour during adaptive walking in older adults. Area of first fixation was used to quantify the spatial location of participants’ initial gaze allocation following movement onset, with lower scores during dual-task conditions indicating a preference for more proximal visual information as older adults begin walking. This proximal shift may reflect greater reliance on visual information concerning the immediately upcoming steps, with less attention directed towards farther areas of the route that could inform subsequent stepping actions. One possible explanation is that the additional attentional demands imposed by the cognitive task limit the resources available for visually sampling and retaining information about the upcoming route, thereby constraining anticipatory gait planning (Ellmers et al., 2016; Ellmers et al., 2020). Compared with single-task walking, participants then went on to perform more transitions towards irrelevant areas and spent longer fixating irrelevant locations during dual-task adaptive walking. This aligns with Ellmers et al. (2016), who reported increased task-irrelevant fixations under cognitive load in young adults, and Russo et al. (2025), who found that dual-tasking reduced proactive visual search towards future stepping targets. Together, these findings suggest that adding a cognitive task reduces participants’ ability to maintain visual attention on areas directly relevant to the walking task, potentially reducing the efficiency of visual information acquisition.

The present findings support previous evidence demonstrating that adaptive walking requires increased cognitive resources and that older adults are vulnerable to dual-task interference (Yogev-Seligmann et al., 2008; Hall et al., 2011; Li et al., 2018; Cocks et al., 2025). This study also replicated the specific dual-task related increase in cross-steps and stepping errors during adaptive walking previously found in older adults (Cocks et al., 2025). When attentional resources are simultaneously allocated to a secondary cognitive task, performance decrements may emerge in both cognitive and motor domains. When attentional resources were allocated to a secondary cognitive task, cross-step rate increased despite participants reporting lower levels of conscious movement processing. This suggests that the increased use of cross-stepping under dual-task conditions may represent a more reactive stepping response. This interpretation is supported by the accompanying gaze behaviour, with dual-tasking associated with more irrelevant fixations and transitions, particularly in the absence of previewing. Reduced task-relevant visual sampling may limit the effective acquisition of information about upcoming stepping constraints and, consequently, the opportunity to use this information proactively to plan subsequent foot placements. Older adults may therefore become more reliant on reactive cross-stepping when cognitive loading interferes with the visual information gathering required for feedforward control of adaptive gait.

### 4.2 Previewing effects on gaze and gait

Following previewing participants made significantly fewer irrelevant fixations and demonstrated greater forward transition sizes, across single- and dual-task conditions, indicating more efficient visual sampling strategy regardless of cognitive loading. Consistent with Curzon-Jones and Hollands (2018), these findings indicate that advance route information modifies subsequent visual sampling during adaptive locomotion and extends this work by demonstrating that these changes are retained under concurrent cognitive loading. Previewing was also associated with significantly lower anxiety and reduced conscious movement processing, with Significant Task × Preview interactions indicating that these were most pronounced under dual-task conditions. These changes were accompanied by improved gait performance (reduced stepping errors and the use of cross-stepping) and attenuation of the detrimental effects of dual-tasking on walking speed. Although no Task × Preview interactions were observed for gaze outcomes, this does not necessarily imply that previewing related gaze changes fail to contribute to the gait benefits observed under dual-task conditions. In single-task walking participants may have had sufficient attentional capacity to meet the demands of adaptive gait, leaving limited scope for previewing-related gaze changes to further improve gait. Under cognitive load, however, finite central processing resources must be shared between concurrent task demands (Tombu & Jolicœur, 2003), potentially increasing the value of visual information acquired in advance. Previewing may therefore reduce the online visual sampling and route planning demands (Curzon-Jones & Hollands, 2018), leaving greater attentional resources available for step control. This interpretation is consistent with evidence that cognitive dual-tasking diverts attentional resources away from concurrent visuomotor processes, disrupting visual sampling and adaptive gait performance (Ellmers et al., 2016; Russo et al., 2025), while reduced previewing of future stepping constraints is associated with greater reliance on online visual control during adaptive gait (Ellmers & Young, 2019), although further research is needed to confirm this mechanism.

Previous research has demonstrated that visual information acquired before and during locomotion aids feedforward planning of future foot placements and adaptive gait adjustments (Chapman & Hollands, 2006; Patla & Vickers, 2003; Matthis et al., 2018). The present findings extend this literature by demonstrating that the benefits of previewing persist for older adults even when attentional resources are challenged by a cognitive dual-task. Older adults typically show reduced attentional capacity (McAvinue et al., 2012) and require more time to generate safe stepping adjustments (Boisgontier., 2013) particularly when negotiating obstacles or unpredictable stepping targets (Caetano et al., 2016). Previewing may therefore provide older adults with the additional time needed for explicit learning to form a stable internal representation of the route before movement begins (Verneau et al., 2014). Such route encoding may allow older adults to pre-plan movement, reducing the need for rapid online adjustments that typically challenge attentional capacity. Such a mechanism is plausible given evidence that, when balance is threatened, older adults allocate attention towards movement processes, threats to balance, and worry-related thoughts, all of which may compete for cognitive resources required for gait control (Ellmers et al., 2020). This may reduce the cognitive load associated with online visual search, freeing attentional resources for step control and thereby attenuating dual-task interference that previously resulted in risky stepping behaviours as a last resort. By decreasing reliance on reactive gait adaptations, route encoding may compensate for age-related declines in obstacle avoidance performance and the slower response times observed when older adults encounter unexpected obstacles during walking (Weerdesteyn et al., 2005).

Route previewing has previously been shown to alter subsequent gaze behaviour and improve stepping safety during adaptive walking (Curzon-Jones & Hollands, 2018). In the current study previewing also attenuated the cognitive dual-task costs observed during adaptive walking. Older adults showed smaller decrements in serial-subtraction performance when they had previewed the route, suggesting that acquiring visual information before movement initiation reduced the attentional demands associated with route negotiation. These cognitive dual-task cost results provide complementary evidence for the potential mechanism described earlier. Whereby, given time to preview the route older adults are able to improve stepping safety by reducing the cognitive demands associated with adaptive gait control. Together, these findings provide the first evidence that route previewing can mitigate both gaze and gait dual-task costs in older adults, even when attentional resources are constrained.

### 4.5 Limitations

Order effects represent an additional limitation. All participants completed the previewing conditions after the non-preview conditions, meaning that improvements observed in the previewing trials may partially reflect familiarity with the walking routes rather than previewing alone. Although the manipulation checks confirmed increased cognitive load during dual-task walking and no increase in anxiety during previewing, the fixed order prevents full separation of previewing effects from potential learning effects. Future studies should counterbalance condition order to minimise this. Although stepping errors were quantified as the total number of incorrect target contacts per trial, this approach does not distinguish between qualitatively different error types. Multiple errors resulting from following an incorrect route may not necessarily reflect greater gait instability than a single isolated foot-placement error, despite producing a substantially higher error count. Future work should therefore differentiate route-selection errors from foot-placement errors, allowing these behaviours to be analysed independently and providing a more meaningful assessment of adaptive gait performance.

### 4.6 Conclusion

The present study investigated whether route previewing could attenuate the negative effects of cognitive dual-tasking on adaptive walking in healthy older adults. Consistent with previous literature, and our first hypothesis, dual-tasking altered adaptive gait performance, resulting in increased stepping errors, increased proportion of cross-steps, reduced gait speed, and greater cognitive dual-task costs. Importantly, these changes were reduced when participants were provided with an opportunity to preview the walking route before movement initiation in line with the third hypothesis. The findings suggest that the possible negative effects of cognitive loading on adaptive walking are not fixed and may be attenuated if some of the visual-motor planning demands are addressed before walking begins. While demonstrating the effectiveness of previewing as a potential intervention, the findings highlight the extent to which cognitive load affects adaptive walking performance and supports the need to consider attentional demands within falls assessment, prevention and rehabilitation.

## Supporting information

Supplemental Results Tables

## 5. Acknowledgements

The authors would like to thank MSc students Emma Vincent, Yi Sum Agatha Siu, Sy Yue, Giuditta Simio, Yasser Bader, and Owen Lin for their assistance with data collection and the Brunel Older Adults Reference Group members for their participation.

## Data Availability

All data produced are available online via Zenodo.

https://doi.org/10.5281/zenodo.23108481

## Footnotes

1 Whereby the cross-stepping outcome was the primary measure as desired by the first hypothesis; we originally planned repeated measures ANOVA, with cross-stepping as the primary outcome of interest to elicit a medium-size effect withing the sample. Subsequently, we opted for linear mixed effects modelling to better accommodate potential incomplete observations. Therefore, our power calculation should be considered an approximate recruitment target. This document: https://doi.org/10.5281/zenodo.23046168 outlines any changes to the methods or analysis previously registered.

