## Supplemental Results Tables for "The effects of route previewing on adaptive gait and gaze behaviour under cognitive dual-task conditions in older adults"

**Supplementary Data**

Gait Outcomes by Condition, Mean (SD)

| **Outcome** | **STNP** | **STP** | **DTNP** | **DTP** |
|---|---|---|---|---|
| Speed (cm/s) | 84.3 (15.4) | 91.8 (16.5) | 60.7 (15.9) | 73.1 (20.3) |
| Stride length (cm) | 95.5 (15.9) | 98.5 (14.7) | 86.2 (14.9) | 91.0 (15.9) |
| Total double support (%) | 32.5 (4.2) | 31.7 (4.2) | 37.7 (5.6) | 35.1 (5.4) |
| Cross-step proportion (Cross-steps/total steps) | 0.0 (0.0) | 0.0 (0.0) | 0.1 (0.1) | 0.0 (0.0) |
| Stepping errors (n) | 0.0 (0.0) | 0.0 (0.3) | 0.9 (0.9) | 0.1(0.2) |

Dual-task Cost Outcomes by Condition, Mean (SD)

| **Outcome** | **DTNP** | **DTP** |
|---|---|---|
| Dual-task cost speed (%) | 27.6 (15.6) | -23.9 (34.7) |
| Dual-task cost step length (%) | 9.54 (8.59) | -5.93 (7.90) |
| Dual-task cost cognitive (%) | 25.1 (31.9) | -23.1 (43.2) |
